# Cytokines as Potential Novel Therapeutic Targets in Severe Inflammatory Cardiomyopathy

**DOI:** 10.1101/2023.07.27.23293253

**Authors:** Phillip Suwalski, Ainoosh Golpour, January Weiner, Nicolas Musigk, Felix Balzer, Niklas Giesa, Ali Amr, Johannes Trebing, Farbod Sedaghat-Hamedani, Benjamin Meder, Dieter Beule, Ulf Landmesser, Bettina Heidecker

**Author notes:** Corresponding author: Bettina Heidecker, MD, Charité Universitaetsmedizin Berlin, Campus Benjamin Franklin, Department of Cardiology. These authors contributed equally (Phillip Suwalski, Ainoosh Golpour).

## Abstract

**Background:** Despite currently available state-of-the art therapies, a substantial proportion of patients with inflammatory cardiomyopathy progresses to advanced heart failure. There is an urgent need for novel therapies to improve outcomes. We hypothesized that elevated cytokine levels in inflammatory cardiomyopathy may lead to cardiac injury and that specific cytokines are associated with severely decreased left ventricular function consequently, thereby suggesting their potential as therapeutic targets.

**Methods and Results:** Blood samples collected from 529 patients at 2 registries were investigated. First, in a derivation cohort of inflammatory cardiomyopathy from our medical center (n=63), we discovered cytokines that correlate inversely with severely decreased left ventricular ejection fraction (LVEF). We confirmed reproducibility of our results in an independent cohort from a national registry (n=425) and to some degree generalizability in a small cohort of idiopathic dilated cardiomyopathy (IDCM, n=41). In total, we identified 82 cytokines associated with severely decreased LVEF (FDR < 0.05); a small portion had been previously proposed as therapeutic targets, while others emerged as novel discoveries. Finally, real-world data from electronic medical records further indicated the potential of inhibitors targeting cytokines of interest to confer a cardioprotective effect.

**Conclusions:** We identified 82 cytokines associated with severe inflammatory cardiomyopathy. Our data were highly significant, reproducible, and generalizable to IDCM. The fact that some of the cytokines had been suggested as potential targets in prior literature supports validity and plausibility of our data. Given that inhibition of cytokines is technically feasible, the identified proteins are compelling potential novel therapeutic targets.

*Trial registration number: ClinicalTrials.gov Identifier: NCT04265040, NCT02187263*

**VISUAL ABSTRACT:** 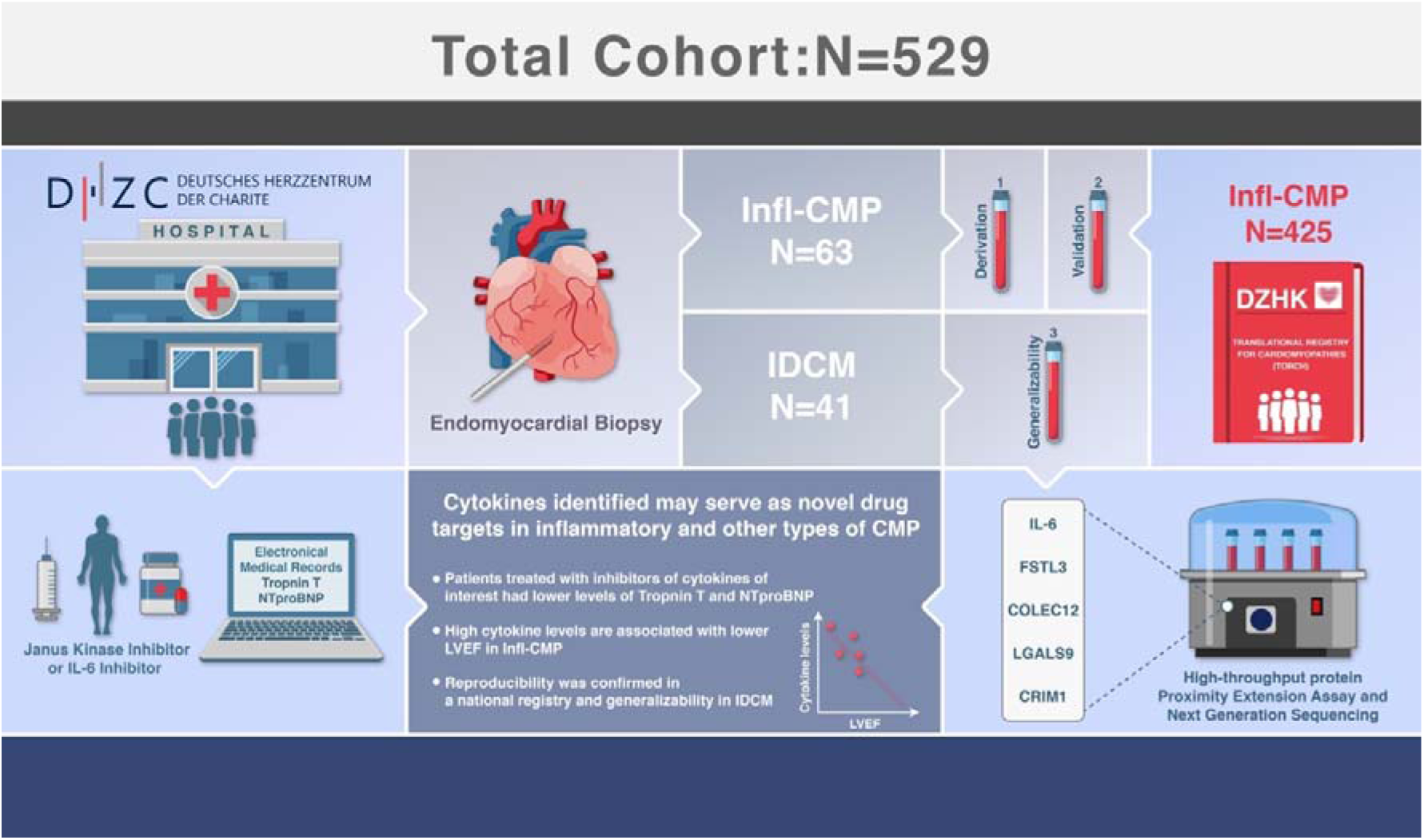

## INTRODUCTION

Inflammatory cardiomyopathy, including myocarditis, is one of the leading causes of sudden cardiac death (SCD) in young adults, with autopsy studies demonstrating that myocarditis is is one of the leading causes of SCD^1^. Despite being a significant public health concern with potentially devastating outcomes, the pathophysiology of inflammatory cardiomyopathy is not well understood, and the condition is often underdiagnosed^2^, leading to suboptimal treatment outcomes for many patients^3^.

The development of drugs for inflammatory cardiomyopathy has been stagnant for more than two decades^4^, with steroids remaining a mainstay of therapy for severe cases^5^. However, steroids are a non-specific treatment that can have various side effects^5^. Despite available treatments, a considerable number of patients initially diagnosed with complicated inflammatory cardiomyopathy progress to chronic heart failure, enduring additional cardiovascular complications^6^.

A recent study following patients with virus-negative chronic inflammatory cardiomyopathy over 20 years found that patients treated with prednisone and azathioprine had better prognostic outcomes than their counterparts, who were treated with standard heart failure therapy alone^7^. Immunosuppressive therapy improved cardiovascular outcomes, including a significantly lower risk of cardiovascular death and heart transplantation, improvement in left ventricular ejection fraction (LVEF) and a lower likelihood of needing an ICD^7^. However, not all patients with virus-negative chronic inflammatory cardiomyopathy respond to immunosuppression with prednisone and azathioprin and may worsen clinically consequently^7, 8^.

Further studies are underway to evaluate the effect of immunosuppressive therapy in virus-negative inflammatory cardiomyopathy in randomized, multicenter, double-blind, pla-cebo-controlled trials (TRINITY, ClinicalTrials.gov: NCT05570409; IMPROVE-MC, ClinicalTrials.gov: NCT04654988).

In recent years, there has been a growing interest in the role of inflammation in heart failure of various etiologies. It is increasingly recognized that a systemic inflammatory state, particularly in inflammatory cardiomyopathy, may contribute to a decline in left ventricular ejection fraction^9^. The impact of systemic inflammation on cardiac injury and the role of cytokines has been increasingly reported in the literature^10–14^.

Therefore, we hypothesized that some cytokines in the context of inflammatory cardiomyopathy may lead to myocardial injury and decreased LVEF consequently. Investigating cytokines that are associated with a severe clinical course of inflammatory cardiomyopathy may help to identify novel drug targets that could be blocked with monoclonal antibodies^15, 16^ or potentially removed through filtration or immunoadsorption techniques^17, 18^. Increasing evidence in the literature suggests that this is a reasonable approach, as blocking cytokines, their receptors, or intracellular kinases further downstream of cytokine signaling were successfully applied in autoimmune diseases and other states of systemic inflammation such as inflammatory cardiomyopathy^3^, sepsis^19^, COVID-19^20^, and in cytokine storm after chimeric antigen receptor (CAR)-T cell treatment^21^.

Our derivation cohort consisted exclusively of patients with biopsy-proven inflammatory cardiomyopathy to ascertain the main results of our study in a cohort with a definitive diagnosis of inflammatory cardiomyopathy. The validation cohort also included patients who were diagnosed based on cardiac magnetic resonance imaging (CMR). Patients were divided into a group with severe inflammatory cardiomyopathy (LVEF ≤ 35%) and patients with mild to moderate inflammatory cardiomyopathy (LVEF > 35%). The choice for a cutoff of 35% for LVEF was guided based on the European Society of Cardiology (ESC) guidelines^22^, in which this cutoff is applied as an indicator for clinical severity and risk of severe arrhythmias. In a third step, we evaluated if our findings were broadly applicable to idiopathic dilated cardiomyopathy (IDCM).

After that, a post-hoc analysis was performed on all samples included in this study.

Finally, to evaluate for potential causality of the discovered cytokines, we evaluated if already existing therapeutic inhibitors of our cytokines of interest have a potential cardioprotective effect. For that purpose, we tested our hypothesis using real world data extracted from electronic medical records of patients who received cytokine inhibitors for various indications at our hospital.

## METHODS

### Study participants and recruitment

We investigated samples from 104 patients who were collected in the biobank of the Deutsches Herzzentrum der Charité (DHZC), Campus Benjamin Franklin, Berlin between 2014 and 2021. Endomyocardial biopsies (EMB) were obtained during routine diagnostic workup as indicated by the ESC Consensus Statement^23^ and American Heart Association Guidelines^24^. As recommended, EMBs were obtained in clinical scenarios, in which the results of histology were expected to change management. Coronary artery disease was excluded in all patients via coronary angiography. Blood samples were systematically collected in EDTA plasma tubes at the time of EMB and stored at -80 °C.

Based on the results of histology and comprehensive immunohistochemistry of EMBs, we divided the patients into 2 groups: 1) Patients with inflammatory cardiomyopathy (n=63) and 2) patients with IDCM (n=41) based on the definition of the ESC^23^.

Patients taking immunosuppressive or anti-inflammatory therapy at the time of EMB were excluded to avoid potential confounders.

Based on echocardiographic data, patients were further divided into a group with severe inflammatory cardiomyopathy (LVEF ≤ 35%) and patients with mild-moderate inflammatory cardiomyopathy (LVEF > 35%). The same was applied to patients with IDCM.

### Replication of results in an independent cohort

To evaluate if our results can be replicated in an independent cohort, we investigated blood samples from patients with inflammatory cardiomyopathy collected in a German national registry for cardiomyopathies. This registry contained samples from patients with a diagnosis of inflammatory cardiomyopathy collected within the Translational registry for cardiomyopathies (TORCH) network of the German Center for Cardiovascular Research e.V. (eingetragener Verein) /Deutsches Zentrum für Herzkreislaufforschung (DZHK).

Patients were recruited at cardiovascular centers across Germany within the DZHK network since 2014. Samples were collected in EDTA tubes and stored within one hour after collection at -80 °C.

Similar to the derivation cohort, patients who had received immunosuppression prior to sample collection were excluded from the analyses. Furthermore, patients missing relevant clinical data such as LVEF were excluded. Samples that qualified for our study were derived from 11 German centers.

### Cytokine analysis

For cytokine analysis, EDTA blood from patients was analyzed with the Proximity Extension Assay (PEA) by Olink. In summary, the method is based on the binding of proteins, i.e. cytokines, by antibodies that carry unique oligonucleotide sequences. When two complementary oligonucleotide sequences come close, they hybridize and can be extended by a DNA polymerase to form a complementary strand. The DNA amplicon can then be quantified using real-time PCR or next generation sequencing (NGS). A major advantage of PEA technology is that it has no impact on data quality while enabling biomarker analysis with high multiplex and fast throughput^25^.

Olink’s Explore 384 – “Inflammation” subpanel, a part of the Explore 3072 panel, was used for our analyses. The full list of cytokines included into the performed Olink Explore 384 – Inflammation panel is provided in the supplement file 1 “Olink 384 Inflammation list”.

The preparation of the sample libraries, randomization, quality control and data processing took place in the process standardized by Olink (for more information, please see the website www.olink.com).Data are presented as normalized protein expression (NPX), providing relative quantification between samples. Log-transformed values were used to obtain a normal distribution.

### Statistical analysis

Statistical analysis was conducted using R. The two batches of Olink data were integrated using the olink_normalization function from the OlinkAnalyze package (v. 3.1) and 16 bridging samples were present in both data sets. Cytokine level influence on LVEF was analysed with a linear regression model (R function lm), using the normalized protein levels (NPX) as the predictor variable and LVEF as the response variable, with cohort (derivation / validation) and patient sex as covariates. P-values were corrected for multiple testing using a false discovery rate controlling procedure (Benjamini-Hochberg method)^26^. For random forest analysis, we used the R package randomForest, version 4.7. R markdown documents and scripts used to generate all results and figures in this manuscript and supplementary data are availa-ble from https://github.com/bihealth/manuscript-myocarditis/.

### Hypothesis testing: Evaluation for potential causality of cytokines in electronic medical record (EMR) real-world data

To further test our hypothesis, we sought to investigate a potential causative effect of cytokines on cardiac injury by analyzing laboratory markers of cardiac injury in patients treated with cytokine inhibitors. These data were collected from three rheumatology departments at the Charité Universitätsmedizin, Berlin. Electronic medical records (EMR) data were extracted from the clinical information system for the period from January 2018 to May 2023. The analysis concentrated on patients who were treated with inhibitors specifically targeting the cytokines identified in our study, or the pathways these cytokines influence downstream. This encompassed patients who had been administered IL-6 or Janus Kinase inhibitors, and for whom high-sensitivity troponin T (hs-TnT) and NT-proBNP measurements were obtainable.

Only laboratory values obtained after the initiation of cytokine inhibitors were included in the analysis. From these laboratory values, the median was calculated for each patient. For patients who did not receive any cytokine inhibitors and therefore belonged to the control group of this analysis, the median was calculated from all available measurements.

For each cytokine being investigated, a control group was established at a 1:4 ratio using the nearest matching procedure, identifying comparable individuals who had not undergone treatment with cytokine inhibitors.

Matching criteria included cardiac, rheumatologic, autoimmune comorbidities, and renal insufficiency. Patients with a glomerular filtration rate (GFR) below 30 ml/min/m² were excluded from the analysis, as hs-TnT and NT-proBNP are affected by renal function. Outliers were identified using the boxplot method^27^ and were excluded from both the respective control group and cytokine inhibitor group. Subsequently, the medians of the respective laboratory values were compared using a Kruskal-Wallis test. Additionally, a linear model was computed.

### Data sharing statement

An anonymized dataset can be made available upon request. Access is only granted to academic staff and after signing a data sharing agreement. The code of the statistical evaluation for the statistics program R is published on GitHub.

### Ethics committee approval

This study was reviewed and approved by the ethics committee of DHZC Universitätsmedizin Berlin (EA4/056/20, EA1/187/22) and the Ethikkommission Medizinische Fakultät Heidelberg (S-344/2014). All patients provided their written informed consent.

In the TORCH registry, all participants provided written informed consent, and the study received approval from the ethics committees of the participating study centers. Additionally, a peripheral blood sample was collected from each participant. Standard Operating Procedure (SOP) for quality training and monitoring was regularly conducted, following the methodology described by Schwaneberg et al. (2017).

The data was analyzed by Phillip Suwalksi, Ainoosh Golpour, January Weiner, Nicolas Musigk and Bettina Heidecker.

## Results

### Study population

We enrolled a total of 529 patients (table 1). The derivation cohort was composed of 63 patients who were recruited from the DHZC, while the validation cohort consisted of 425 patients who were enrolled within the TORCH registry of the DZHK.

**Table 1:** Demographic data: Derivation cohort; independent validation cohort; generalizability cohort: Cohort of patients with idiopathic dilated cardiomyopathy, in whom general applicability of the findings was assessed.

|  | **Derivation cohort (n=63)** | **Validation cohort (n=425)** | **Generalizability cohort (n=41)** |
| --- | --- | --- | --- |
| Cardiomyopathy | Inflammatory Cardiomyopathy | Inflammatory Cardiomyopathy | Idiopathic Dilated Cardiomyopathy |
| Registry | DHZC | DZHK | DHZC |
| Sex female (n, %) | 19 (30.2%) | 120 (28.2%) | 13 (31.7%) |
| Age in years (median, IQR) | 54 (36 – 62) | 49 (37 – 58) | 54 (48 – 65) |
| BMI in kg/m<sup>2</sup> (median, IQR) | 27 (25 – 29) | 26 (24 – 30) | 27 (24 – 32) |
| LVEF ≤ 35% (n, %) | 23 (36.5%) | 169 (39.8%) | 17 (41.5%) |
| LVEF (median, IQR) | 45 (31 – 58) | 42 (30 – 55) | 42 (30 – 58) |
| Creatinine in mg/dl (median, IQR) | 0.93 (0.83 – 1.1) | 0.93 (0.82 – 1.1) | 0.94 (0.85 – 1.1) |

The derivation (n=63) and validation (n = 425) cohort contained only patients with inflammatory cardiomyopathies. The third group, referred to as the “Generalizability” cohort, encompassed patients diagnosed with IDCM (n=41).

There were no significant differences among the groups concerning key baseline characteristics such as sex, age, BMI, LVEF, and renal function. Women comprised between 28.2% and 31.7% of the participants across all cohorts. The median age of the participants spanned from 49 to 54 years. Moreover, the median Body Mass Index (BMI) was in the range of 26-27 kg/m², categorizing the participants, on average, as overweight.

The three groups were categorized based on the extent of reduction in LVEF, with patients classified as having either severe LVEF reduction (≤ 35%) or mild to moderate LVEF reduction (>35%). The median LVEF was in the range of 42-45%. Furthermore, the median creatinine levels ranged from 0.93 to 0.94 mg/dl, indicative of normal renal function.

### Cytokine Analysis in the Derivation Cohort

First, we tested the hypothesis that cytokine levels are associated with severe inflammatory cardiomyopathy, defined by an LVEF ≤35%. A linear regression model was employed to analyze the relationship between cytokine levels and LVEF in patients with inflammatory cardiomyopathy.

Following correction for multiple testing, a total of 5 cytokines were found to be significantly associated with LVEF in the derivation cohort of patients with inflammatory cardiomyopathy (DHZC, n= 63) (FDR < 0.05; Supplemental table 1; Supplemental File 2 (full_results.xlsx). The relationship between relative cytokine concentration and LVEF was found to be inverse. This indicates that higher relative cytokine concentrations are associated with lower LVEF in patients with inflammatory cardiomyopathy.

### Normalization and Cytokine Analysis in Validation cohort

In order to validate the results obtained from the derivation cohort, we utilized patients diagnosed with inflammatory cardiomyopathy from the TORCH registry of the DZHK as a validation cohort. The samples were collected in two batches. The first batch comprised of the derivation and generalizability cohorts, and the second batch included the validation cohort as well as 16 bridging samples. The normalization procedure followed the recommendations of the manufacturer.

The data were normalized using the OlinkAnalize package, version 3.1.0, using the first batch data set as the reference sample and 16 bridging samples.

Results of the normalization process are presented in Supplemental Figure 1 and 2.

Principal component analysis was performed to ensure that no subgroups differentiated the derivation and validation datasets. No significant differences were observed for the first and second principal components between the datasets. As such, the datasets were considered comparable (Supplemental Figure 3 and Figure 4)

Similar to the derivation cohort, a linear regression model was employed to analyze the relationship between cytokine levels and LVEF in the independent validation cohort of patients with inflammatory cardiomyopathy collected in the national registry (TORCH, n= 425). Following correction for multiple testing, a total of 77 cytokines were found to be significantly associated with LVEF (FDR < 0.05; Supplemental table 2; Supplemental File 2 (full_results.xlsx)).

The relationship between relative cytokine concentration and LVEF was again found to be inverse. The replication in the validation cohort confirms that higher relative cytokine concentrations of the top 10 cytokines identified in the derivation cohort (DHZC) are associated with lower LVEF in patients with inflammatory cardiomyopathy from the independent cohort (TORCH; Supplemental table 3).

### Generalizability of findings

Following the identification of cytokines that showed an inverse relationship with LVEF in patients with inflammatory cardiomyopathy, generalizability of these findings was evaluated in patients with IDCM (DHZC, n=41). A Welch two-sided t-test was used to compare the groups with severe vs mild to moderately reduced LVEF in IDCM. Similar to the derivation and validation cohort, there was a trend for top cytokines of both cohorts being associated with more severely reduced LVEF in IDCM as well (Figure 1). After correction for multiple testing, no cytokines were significant. However, there was a significant correlation between the effect sizes of the derivation and generalizability cohort (Pearson’s correlation coefficient = 0.29).

**Figure 1:**
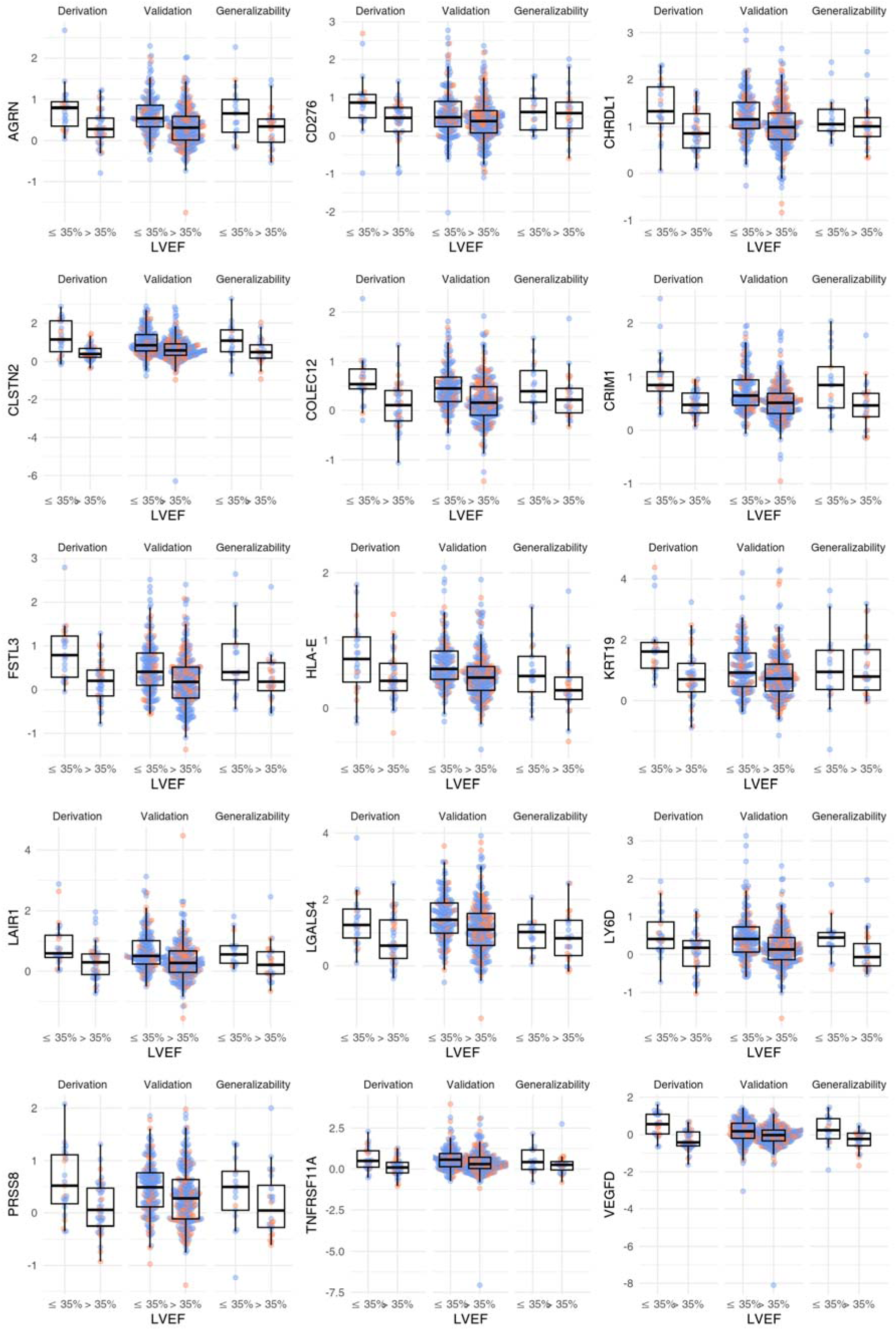
Relative cytokine concentrations in patients with inflammatory cardiomyopathy in both derivation and validation cohort (n = 488). The patients were grouped according to cohort and left ventricular ejection fraction (LVEF). The selected cytokines are the top 15 cytokines identified in the derivation cohort. Blue dots correspond to male patients, red dots correspond to female patients. We tested whether there was a significant interaction between LVEF and sex. We did not find any proteins which showed both, a significant main effect of LVEF and a significant interaction between LVEF and sex, neither in the derivation cohort nor in the validation cohort. Association between LVEF and cytokine levels was significant for all cytokines shown at FDR < 0.001.

Complete values for all analyzed cytokines can be found in the Supplemental File (Supple-mental File 2 (full_results.xlsx)).

### Post hoc analysis for all patients with inflammatory cardiomyopathy

After normalization, the relative cytokine values of all 488 patients with inflammatory cardiomyopathy were collectively analyzed and compared to each other with regards to their ability to distinguish between severe and mildly reduced LVEF categories. Covariables sex and dataset of origin were used in the analysis. A total of 82 proteins were found to be significant after correction for multiple testing. The top values for the top cytokines of our post hoc analysis are presented in table 2 (complete available in Supplemental File (full_results.xlsx).

**Table 2:** Top 10 cytokines associated with left ventricular ejection fraction (LVEF): Cy-tokines were derived from the derivation and validation cohorts when analyzed together (n = 488). Higher relative cytokine concentrations are associated with lower LVEF values.

| Cytokine | Estimate | r<sup>2</sup> | p value | FDR |
| --- | --- | --- | --- | --- |
| COLEC12 | -12.58 | 0.1762 | < 2e-16 | < 2e-16 |
| PLAUR | -11.25 | 0.163 | < 2e-16 | 4.2e-15 |
| AGRN | -11.2 | 0.1549 | < 2e-16 | 4.6e-14 |
| WNT9A | -10.03 | 0.1492 | 6.5e-16 | 2.4e-13 |
| LAIR1 | -8.063 | 0.144 | 2.9e-15 | 1.0e-12 |
| LILRB4 | -8.768 | 0.1439 | 3.0e-15 | 1.1e-12 |
| FABP1 | -4.191 | 0.1416 | 5.8e-15 | 2.1e-12 |
| CRIM1 | -13.45 | 0.1403 | 8.4e-15 | 3.0e-12 |
| CCL3 | -6.865 | 0.1389 | 1.3e-14 | 4.5e-12 |
| FSTL3 | -8.187 | 0.1324 | 8.1e-14 | 2.9e-11 |

### EMR Data Evaluation

We screened 18,566 patient records from three rheumatology departments of Charité University Berlin to test our hypothesis in EMR data. Among these, hs-TnT and/or NT-proBNP measurements were available for 8,767 patients. Within this patient pool, 348 individuals had been administered the cytokine inhibitors of interest. These included IL-6 inhibitors and janus kinase inhibitors.

For the other cytokines identified in our study, there are either no specific inhibitors presently in clinical use, or the number of patients treated was insufficient to permit robust statistical analysis.

Out of this subset, 190 patients had hs-TnT and/or NT-proBNP measurements recorded post-administration of the cytokine inhibitors. After excluding statistical outliers, the final analysis consisted of a total of 159 patients.

#### NT-proBNP

Treatment with IL-6 receptor inhibitors (n=69) resulted in a notable reduction in median NT-proBNP levels by 136 ng/L (p < 0.001). Similarly, in the group receiving janus kinase inhibition (n=90) the median NT-proBNP level was significantly reduced by approximately 170.5 ng/L (p < 0.001). The results are presented in Figure 2.

**Figure 2:**
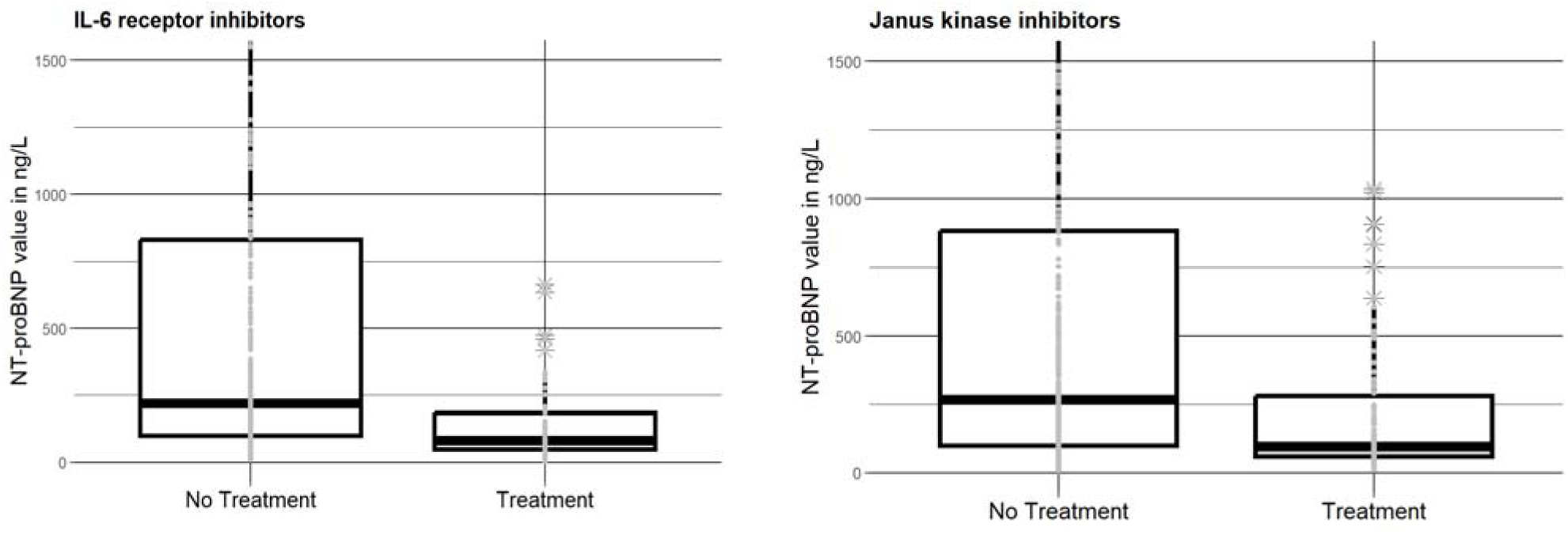
Comparison of NT-proBNP values in ng/L between patients who were prescribed IL-6 receptor inhibitors or janus kinase inhibitors with matched controls not taking those drugs. IL-6 receptor inhibitor or janus kinase inhibitor intake was associated with lower NT-proBNP values.

#### Hs-TnT

In the group receiving janus kinase inhibitors (n=52) a significant decrease in median hs-TnT levels by 9 ng/L (p = 0.001) was detectable. The results are presented in Figure 3.

**Figure 3:**
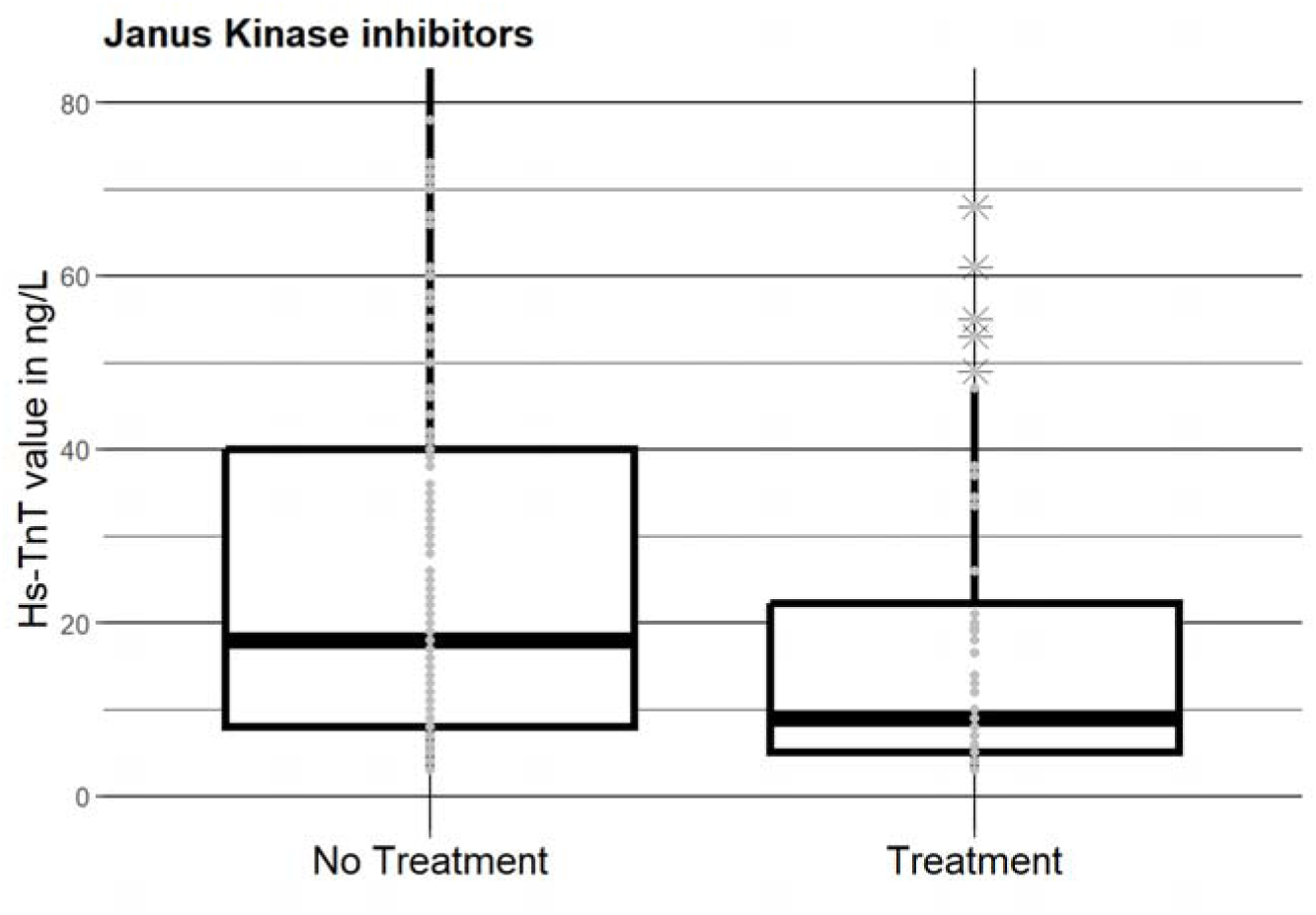
Janus kinase inhibitor use was linked to lower high-sensitivity Troponin T (hs-TnT, ng/L) values when compared to matched controls not taking this drug.

## DISCUSSION

This study aimed to fill a crucial clinical gap in the treatment of inflammatory cardiomyopathy, by exploring new potential therapeutic targets for patients who do not respond to existing, advanced therapies. We hypothesized that some cytokines released in the context of inflam-matory cardiomyopathy may lead to myocardial injury and decreased LVEF as a consequence. Therefore, we investigated the association of cytokines with LVEF. Our investigation proved our hypothesis that there was an inverse correlation of specific cytokines with LVEF.

We first identified our findings within a derivation cohort from our medical center (n=63). Then, we replicated our results in an independent validation cohort (n=425), which encompassed samples gathered from 11 different cardiovascular centers across Germany, as part of the national TORCH registry. Furthermore, testing of our results in patients with IDCM (n=41) suggested generalizability of our findings. A post-hoc analysis further confirmed the robustness of our data. Real-world data from our EMR system, specifically from patients treated with inhibitors targeting the cytokines identified in our study or the pathways they influence downstream, suggested a potential cardioprotective effect of such specific cytokine inhibition. These data support the validity of our hypothesis and findings.

All three cohorts, including derivation, validation, and generalizability cohort, demonstrated a balanced distribution of key baseline parameters including sex, age, BMI, LVEF, and renal function.

The sex distribution, with approximately one third of the participants being women is in agreement with prior literature^3, 10, 13, 28^. Based on the median BMI, a large part of the patients was overweight. This is an observation that aligns with findings from prior work on inflam-matory cardiomyopathy^28^. One may speculate that proinflammatory characteristics of adipose tissue could contribute to a predisposition of developing inflammatory cardiomyopathy^28^.

A considerable proportion of patients (36.5% in the derivation cohort, 39.8% in the validation cohort and 41.5% in the generalizability cohort) exhibited severely compromised left ventricular function, ensuring a balanced representation of the degree of illness. Kidney function was overall preserved in all cohorts.

According to ESC guidelines^22^, patients with LVEF ≤35% received more angiotensin-receptor blockers (ARBs) and diuretics compared to those with a higher LVEF. If this disparity in medication regimen influenced our study outcome, it likely concealed the observed association between specific cytokines and low LVEF, given that ARBs are known to possess anti-inflammatory properties^29, 30^.

Our investigation confirmed the hypothesis that specific cytokines correlate with severe inflammatory cardiomyopathy as defined by severely reduced LVEF (≤35%). This correlation was inverse, highly significant, and reproducible in a large independent set of samples that were collected at 11 different sites in Germany in a national registry. Furthermore, our data suggested that results may be generalizable to IDCM.

Several of the cytokines identified had been suggested or evaluated as potential therapeutic targets in inflammatory cardiomyopathy previously, supporting validity and plausibility of our results.

Similar to prior studies in patients with heart failure overall^31–34^, higher serum levels of IL6 or CRIM1 were associated with more severe forms of inflammatory cardiomyopathy in our study. Importantly, it had been shown previously that in IL6 deficient mice, prevalence and severity of myocarditis were reduced in the absence of IL6^31^. In myocarditis, IL6 is produced by monocytes infiltrating the myocardium and differentiating into inflammatory macrophages, contributing to tissue degradation and T cell activation^3^.

IL6 inhibitors tocilizumab and sarilumab have been administered successfully to patients with severe COVID-19 to mitigate the overactive cytokine response, improving survival^35^.

Janus kinase inhibitors block downstream signaling of inflammatory pathways of IL6 and other cytokines. Investigation of our current EMR data demonstrating lower NT-proBNP and hs-TnT levels in patients receiving IL6 or janus kinase inhibitors is in agreement with existing literature suggesting that this drug classes reduce cardiac inflammation and injury^31, 36, 37^.

Our study identified numerous novel cytokines that had either not been previously associated with severe inflammatory cardiomyopathy or had only been minimally explored within this context. These included COLEC 12 - a scavenger receptor that plays a role in host defense, LAIR1 – a protein expressed on human leukocytes and LILRB4 – a leukocyte immunoglobu-lin-like receptor (http://www.genenames.org/).

FSTL3 was recently identified as relevant prognostic marker for major adverse cardiovascular events in stable coronary artery disease. Higher levels of FSTL3 were associated with poor outcomes^38^. Also in the current study, higher blood levels of FSTL3 were associated with severe inflammatory cardiomyopathy.

LGALS9 is a protein that modulates cell-cell and cell-matrix interactions. It has been shown to promote innate immune cell activation by potentiating or synergizing toll-like receptor signaling^39^. Also, LGALS9 has been associated with the severity of myocarditis in mice infected with Coxsackievirus B3 and to have an immunomodulatory effect when administered in experimental myocarditis^40^.

Our findings have important implications for future treatment of patients with inflammatory cardiomyopathy, who clinically deteriorate despite currently available state-of-the art medical therapy. Cytokines are feasible therapy targets as prior research has shown. Blocking of cytokines, their receptors, or intracellular kinases downstream of cytokine signaling has been successfully applied^4, 16, 19, 20, 21, 22^. Also, removal through immunoadsorption or -filtration may be applied. Drug-repurposing of already existing drugs that target the cytokines of inter-est such as Ixekizumab and Secukinumab to neutralize IL17A could be another approach that will be tested in future experiments.

## LIMITATIONS

Our study identified numerous potential novel therapy targets in inflammatory cardiomyopathy that may be of value in patients, whose clinical condition worsens as they do not respond to current optimal medical therapy. To identify the optimal targets among those reported, further experiments on cardiomyocytes in cell culture as well as animal experiments will be necessary to evaluate for causality and potential side effects of inhibition. The cohort of IDCM was small. Generalizability of our findings will have to be tested in a larger cohort of IDCM.

## CONCLUSIONS

We tested the hypothesis that there is excess of specific cytokines in severe inflammatory cardiomyopathy, which may lead to cardiac injury and decreased LVEF as a result. Our investigation confirmed that 82 cytokines were elevated in patients with severe inflammatory cardiomyopathy. Furthermore, elevation of cytokine levels correlated inversely with LVEF. The data were not only statistically significant and reproducible, but also to a certain extent applicable to cases of IDCM.

Some of these cytokines had been previously reported as potential therapy targets in inflam-matory cardiomyopathy and heart failure in general, supporting validity and biological plausibility of our data. In addition, we discovered numerous novel cytokines of interest that have the potential to be used as therapeutic targets through binding and neutralizing agents, filtra-tion, immunoadsorption or drug repurposing of already existing drugs in patients, who clinically deteriorate despite current state-of-the art therapies.

## Supporting information

Cytokine list OLINK 384

Full results

## ACKNOWLEDGEMENTS

We thank all TORCH investigators and staff, Andrea Heuberger, Michele Violano, and Xiaomin Wang for their help with sample collection and sample processing.

