## Supplement for "Cytokines as Potential Novel Therapeutic Targets in Severe Inflammatory Cardiomyopathy"

**Supplemental Table 1:** Derivation cohort (n=63): Top 10 cytokines associated with left ventricular ejection fraction (LVEF). Higher relative cytokine concentrations are associated with lower LVEF values. This inverse relationship with LVEF was significant for 5 cytokines (FDR < 0.05). Estimate, the coefficient estimate from the linear regression model; $r^2$, coefficient of determination ($r^2$) value for the model; P-value, raw p value for the model; FDR, false discovery rate calculated using Benjamini-Hochberg correction.

| Cytokine | Estimate | r2 | P-value | FDR |
|---|---|---|---|---|
| VEGFD | -15,418 | 0,421638 | 5,47E-08 | 2,01E-05 |
| CLSTN2 | -12,3104 | 0,315359 | 1,01E-05 | 0,003696 |
| FSTL3 | -14,615 | 0,313893 | 1,08E-05 | 0,00394 |
| KRT19 | -8,64163 | 0,31284 | 1,13E-05 | 0,004121 |
| CRIM1 | -20,57 | 0,287229 | 3,54E-05 | 0,012901 |
| COLEC12 | -14,5942 | 0,244778 | 0,000219 | 0,079625 |
| CHRDL1 | -13,612 | 0,228297 | 0,000436 | 0,15771 |
| LAIR1 | -10,3162 | 0,222985 | 0,000542 | 0,195762 |
| PRSS8 | -12,4807 | 0,22271 | 0,000548 | 0,197437 |
| LY6D | -11,7192 | 0,217577 | 0,000677 | 0,243021 |

**Supplemental Table 2:** Validation cohort (n=452): Similar to the derivation cohort, there is an inverse relationship between cytokine concentrations and left ventricular ejection fraction (LVEF) with high cytokine levels being associated with lower LVEF. There were 77 cytokines significantly associated with LVEF (FDR < 0.05). Estimate, the coefficient estimate from the linear regression model; $r^2$, coefficient of determination ($r^2$) value for the model; P-value, raw p value for the model; FDR, false discovery rate calculated using Benjamini-Hochberg correction.

| Cytokine | Estimate | r2 | P-value | FDR |
|---|---|---|---|---|
| COLEC12 | -12,1741 | 0,164675 | 4,46E-16 | 1,64E-13 |
| PLAUR | -11,1866 | 0,163439 | 6,12E-16 | 2,24E-13 |
| CCL3 | -7,14421 | 0,152285 | 1,04E-14 | 3,81E-12 |
| LILRB4 | -9,01034 | 0,151545 | 1,25E-14 | 4,58E-12 |
| CXCL17 | -5,54511 | 0,149364 | 2,18E-14 | 7,92E-12 |
| AGRN | -10,9161 | 0,148426 | 2,75E-14 | 1E-11 |
| WNT9A | -9,78385 | 0,141907 | 1,41E-13 | 5,11E-11 |
| CD4 | -11,4165 | 0,137435 | 4,31E-13 | 1,56E-10 |
| FABP1 | -4,20844 | 0,136129 | 5,96E-13 | 2,15E-10 |
| LAIR1 | -7,74702 | 0,133987 | 1,01E-12 | 3,64E-10 |
| LGALS9 | -8,96591 | 0,123841 | 1,24E-11 | 4,44E-09 |
| CRIM1 | -12,4333 | 0,122375 | 1,78E-11 | 6,35E-09 |
| TNFSF13 | -10,0253 | 0,121528 | 2,19E-11 | 7,79E-09 |
| FSTL3 | -7,4228 | 0,113642 | 1,5E-10 | 5,33E-08 |
| CCL7 | -4,43945 | 0,109818 | 3,8E-10 | 1,35E-07 |
| TGFA | -7,94094 | 0,109175 | 4,44E-10 | 1,57E-07 |
| CLSTN2 | -6,20831 | 0,107495 | 6,67E-10 | 2,35E-07 |
| HGF | -4,49271 | 0,104511 | 1,37E-09 | 4,82E-07 |
| ANGPTL4 | -6,68144 | 0,104232 | 1,47E-09 | 5,14E-07 |
| SPON1 | -7,40211 | 0,103125 | 1,92E-09 | 6,69E-07 |
| IL17D | -9,61421 | 0,101249 | 3,01E-09 | 1,05E-06 |
| LRRN1 | 7,205829 | 0,101132 | 3,1E-09 | 1,08E-06 |
| LY6D | -7,43169 | 0,098996 | 5,18E-09 | 1,79E-06 |
| PON3 | 11,39412 | 0,095766 | 1,13E-08 | 3,88E-06 |
| HLA-E | -11,4467 | 0,095722 | 1,14E-08 | 3,91E-06 |
| SPINK4 | -4,40881 | 0,095501 | 1,2E-08 | 4,11E-06 |
| LGALS4 | -5,27358 | 0,09436 | 1,58E-08 | 5,39E-06 |
| IL6 | -2,99671 | 0,093813 | 1,8E-08 | 6,13E-06 |
| NPPC | -4,98094 | 0,093211 | 2,08E-08 | 7,06E-06 |
| CHRDL1 | -7,91593 | 0,092942 | 2,21E-08 | 7,51E-06 |
| TREM2 | -4,77254 | 0,091431 | 3,18E-08 | 1,07E-05 |
| CXCL10 | -4,17866 | 0,088631 | 6,21E-08 | 2,09E-05 |

| Cytokine | Estimate | r2 | P-value | FDR |
|---|---|---|---|---|
| EPO | -3,74666 | 0,087718 | 7,72E-08 | 2,59E-05 |
| CXCL8 | -4,01822 | 0,087691 | 7,77E-08 | 2,6E-05 |
| MATN2 | -9,39644 | 0,086945 | 9,28E-08 | 3,1E-05 |
| CXCL14 | -4,49222 | 0,08555 | 1,29E-07 | 4,31E-05 |
| GAL | 4,65288 | 0,082948 | 2,4E-07 | 7,98E-05 |
| TFF2 | -4,06551 | 0,081606 | 3,31E-07 | 0,000109 |
| PREB | -10,3113 | 0,081157 | 3,68E-07 | 0,000121 |
| SULT2A1 | -4,04533 | 0,078762 | 6,5E-07 | 0,000214 |
| ENPP7 | -3,32201 | 0,077855 | 8,06E-07 | 0,000264 |
| IL15 | -6,78804 | 0,077687 | 8,39E-07 | 0,000274 |
| DNER | 9,589636 | 0,077289 | 9,22E-07 | 0,0003 |
| CXCL9 | -3,22193 | 0,074958 | 1,6E-06 | 0,000521 |
| IL4R | -6,07854 | 0,074123 | 1,95E-06 | 0,000633 |
| REG4 | -4,97668 | 0,073114 | 2,48E-06 | 0,000801 |
| BTN3A2 | -6,09672 | 0,071531 | 3,61E-06 | 0,001162 |
| FST | -5,18705 | 0,071502 | 3,63E-06 | 0,001166 |
| EGLN1 | -3,95482 | 0,07108 | 4,01E-06 | 0,001284 |
| TNF | -5,94492 | 0,070676 | 4,42E-06 | 0,001409 |
| IL1RN | -3,4451 | 0,070089 | 5,08E-06 | 0,001614 |
| NTF3 | -5,07255 | 0,069915 | 5,29E-06 | 0,001677 |
| TNFRSF13B | -6,8624 | 0,069553 | 5,76E-06 | 0,001821 |
| CXADR | -4,82297 | 0,068877 | 6,76E-06 | 0,00213 |
| CD276 | -5,47858 | 0,068871 | 6,77E-06 | 0,00213 |
| TNFRSF11A | -4,2661 | 0,06851 | 7,38E-06 | 0,002308 |
| SMOC2 | -5,99596 | 0,068308 | 7,73E-06 | 0,002413 |
| CCL25 | -4,11859 | 0,067723 | 8,88E-06 | 0,002763 |
| NFASC | -7,98799 | 0,067721 | 8,89E-06 | 0,002763 |
| CCL28 | -3,05271 | 0,067265 | 9,9E-06 | 0,003059 |
| SIGLEC10 | -5,64472 | 0,066844 | 1,09E-05 | 0,003368 |
| PRSS8 | -5,97756 | 0,066519 | 1,18E-05 | 0,003626 |
| LAMA4 | -6,9675 | 0,066442 | 1,2E-05 | 0,00368 |
| PRELP | -8,7097 | 0,065305 | 1,57E-05 | 0,004801 |
| ENPP5 | 5,817113 | 0,063325 | 2,51E-05 | 0,007643 |
| CCL21 | -4,3005 | 0,063229 | 2,57E-05 | 0,007793 |
| TNFRSF4 | -4,64774 | 0,061676 | 3,71E-05 | 0,011215 |
| HLA-DRA | -5,76163 | 0,061424 | 3,94E-05 | 0,011865 |
| CCL23 | -5,004 | 0,061069 | 4,29E-05 | 0,012863 |
| FASLG | 4,476295 | 0,060662 | 4,72E-05 | 0,014115 |
| TPP1 | -5,9269 | 0,059997 | 5,53E-05 | 0,016466 |
| CCL11 | -4,03204 | 0,059247 | 6,6E-05 | 0,019599 |
| CCL13 | -2,60901 | 0,057756 | 9,39E-05 | 0,0278 |
| KRT19 | -3,53618 | 0,057209 | 0,000107 | 0,031541 |
| LIFR | -6,6545 | 0,05658 | 0,000124 | 0,036485 |
| ITM2A | -4,51045 | 0,056553 | 0,000125 | 0,036589 |
| CKAP4 | -5,15816 | 0,056407 | 0,000129 | 0,037749 |

**Supplemental Table 3:** Replication in the validation cohort confirms that higher relative cytokine concentrations of the top 10 cytokines from the derivation cohort are associated with lower LVEF in patients with inflammatory cardiomyopathy. Estimate, the coefficient estimate from the linear regression model; $r^2$, coefficient of determination ($r^2$) value for the model; P-value, raw p value for the model; FDR, false discovery rate calculated using Benjamini-Hochberg correction.

| Cytokine | Estimate | r2 | p value | FDR |
|---|---|---|---|---|
| COLEC12 | -12.17 | 0.1647 | 4.5e-16 | 1.6e-13 |
| LAIR1 | -7.747 | 0.134 | 1.0e-12 | 3.6e-10 |
| CRIM1 | -12.43 | 0.1224 | 1.8e-11 | 6.3e-09 |
| FSTL3 | -7.423 | 0.1136 | 1.5e-10 | 5.3e-08 |
| CLSTN2 | -6.208 | 0.1075 | 6.7e-10 | 2.3e-07 |
| LY6D | -7.432 | 0.099 | 5.2e-09 | 1.8e-06 |
| CHRDL1 | -7.916 | 0.09294 | 2.2e-08 | 7.5e-06 |
| PRSS8 | -5.978 | 0.06652 | 1.2e-05 | 0.0036 |
| KRT19 | -3.536 | 0.05721 | 0.00011 | 0.0315 |
| VEGFD | -3.324 | 0.04488 | 0.00202 | 0.5383 |

**Supplemental Figure 1:** NPX values of all protein assays in the first batch (X axis, derivation and generalizability cohorts) and second batch (Y axis, validation cohort) data sets for the 16 bridging samples. Each dot represents one protein assay in one sample. Left panel, raw data. Right panel, data normalized using the olink_normalization function from the OlinkAnalyze R package. In total, there were 16 samples, 368 assays and 5888 total measurements. Blue line shows a fitted LOESS model.

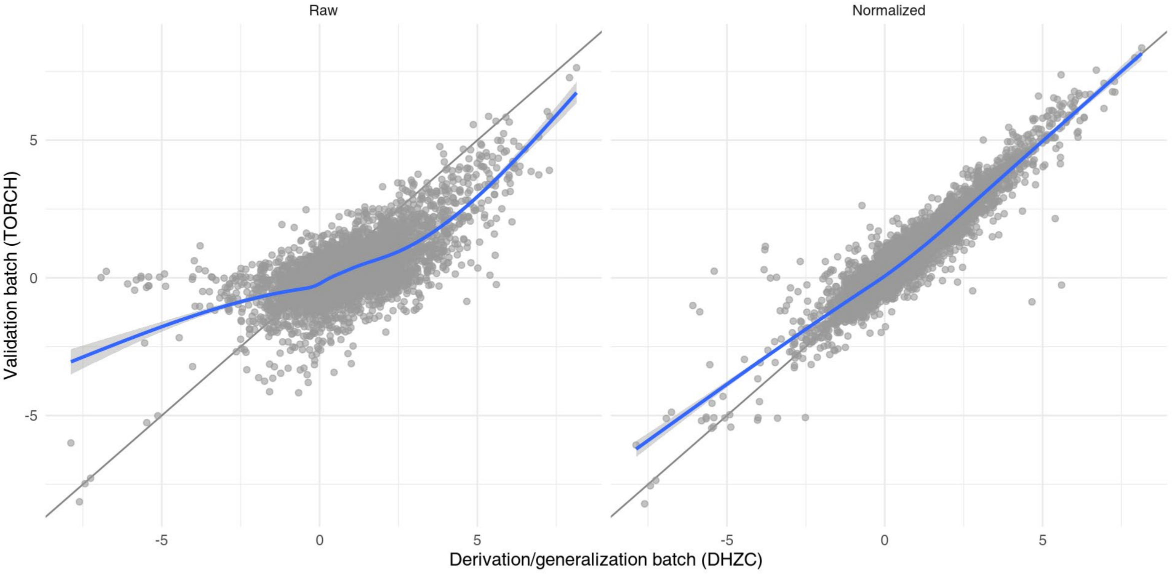

**Supplemental Figure 2:** Comparison of 10 randomly chosen protein assays across the different data sets and different normalizations for the 16 bridging samples. Top: raw data. Bottom: normalized data. In total, 16 unique samples and 640 measurements are shown.

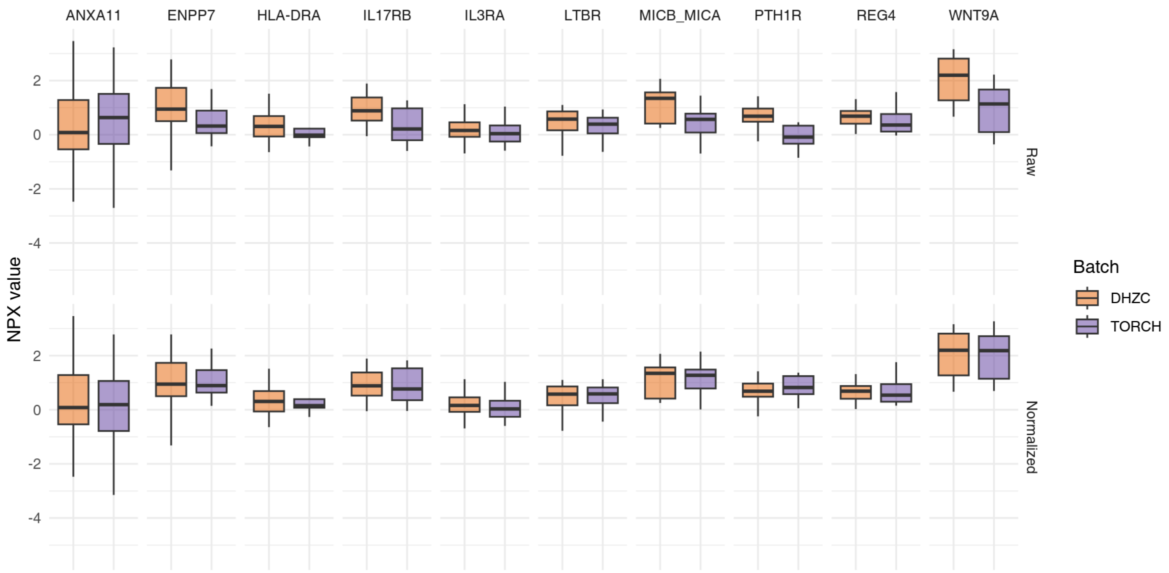

**Supplemental Figure 3:** Principal component analysis of the 529 measurements before normalization (left panel) and the same measurements after normalization (right panel). Each dot represents one sample. The first two principal components are shown in the top row, and the third and fourth principal components are shown in the bottom row. The color of the dots indicates the batch of the sample.

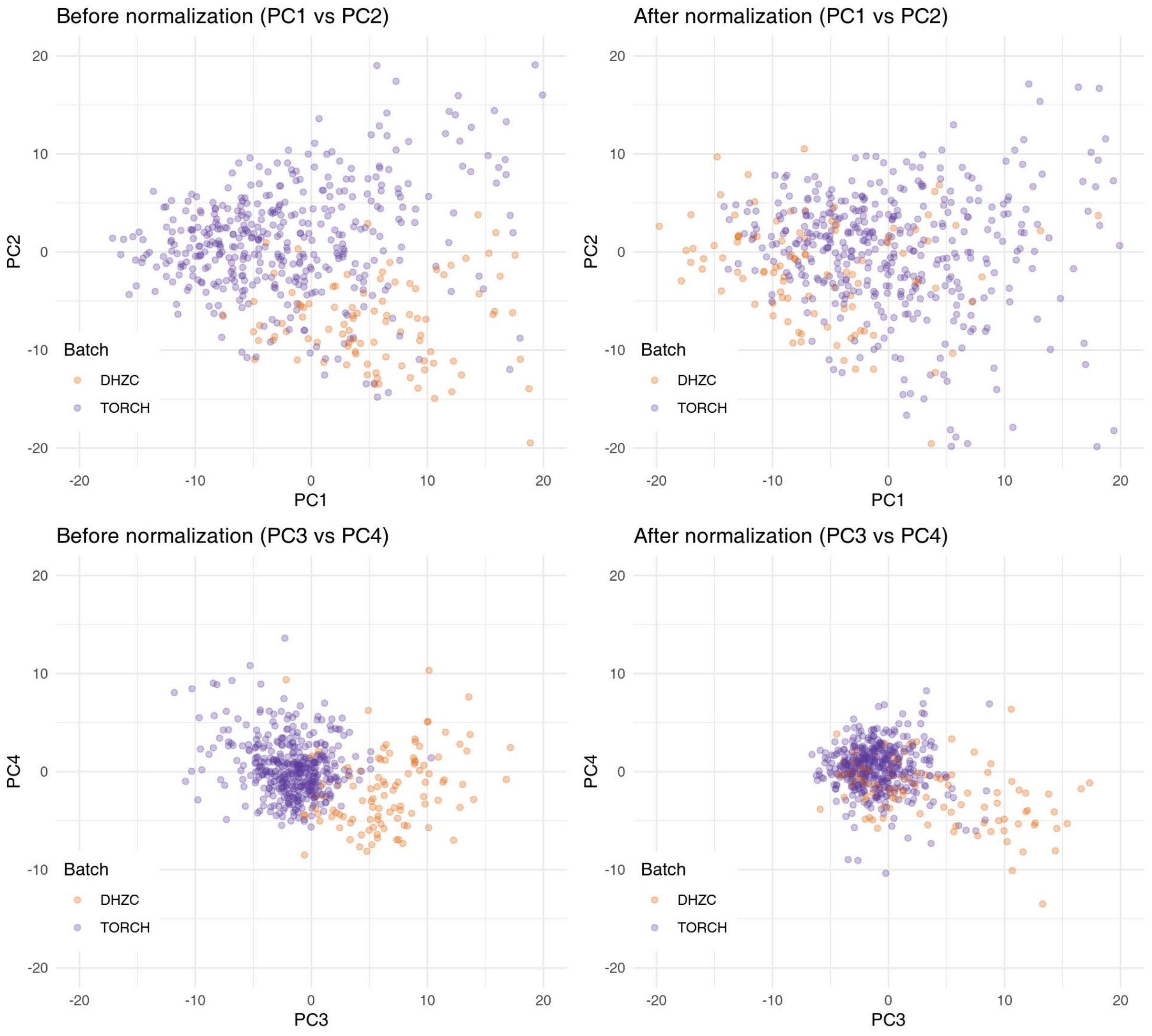

**Supplemental Figure 4**: Scree plot of the principal component analysis of the 529 measurements before normalization (red) and the same measurements after normalization (blue). The x-axis shows the principal components and the y-axis shows the proportion of variance explained by each principal component.

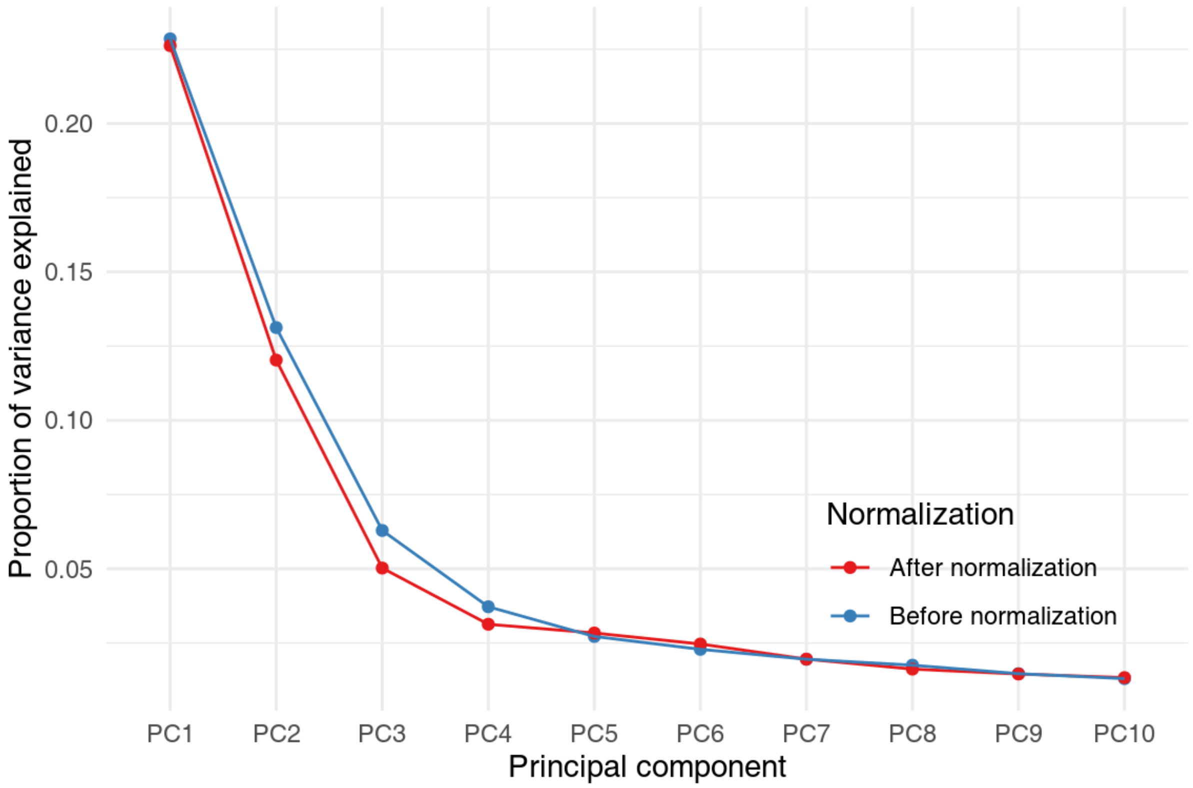
